# Cost of Retracted Articles to the NIH: a Living Analysis

**DOI:** 10.64898/2026.07.04.26357187

**Authors:** Alejandro Sandoval-Lentisco, John P.A. Ioannidis

## Abstract

Retractions attract substantial attention and have become more frequent over time. Retractions reflect the self-correcting nature of science but also wasted resources. The Retraction Watch Database (RWD) includes over 60,000 records. We integrated RWD with NIH funding metadata (RePORTER system). As of July 2026, of the 6,081 U.S. affiliated retracted articles, 1,725 (28.4%) were linked to at least one NIH grant. With a mean attributed cost per retracted NIH-funded article of $255,087 in 2026 dollars, the attributed total cost of NIH-funded retracted research is $440 million in 2026 dollars. Grants associated with retracted papers for which the first or last author of the paper was the principal investigator were awarded $4.03 billion in 2026 dollars. NIH-funded articles take longer to be retracted (mean = 6.3 years) than other US-based articles, which may entail greater downstream implications. NIH funding of retracted authors decreased over the 3 years following retraction, particularly among authors with multiple retractions. We have developed a continuously updated dynamic dashboard (https://sandovallentisco.shinyapps.io/nih-retractions/) for the cost of retractions reflecting NIH-funded work.

## Introduction

Retracted publications are a visible manifestation of how science self-corrects, but they may also signal serious weaknesses in research practices and peer review (*1*). Retracted work, especially when it is due to serious fraudulent practices or major flaws and not just publisher or journal technical errors, may undermine trust in science, distort scientific evidence, and represent a waste of scarce research resources. Although retractions remain rare relative to overall publication volume, their frequency has been climbing for two decades (*2*, *3*). One would wish to estimate the cost of the research work that has been retracted. Previous estimates of the cost of retracted research for the U.S. National Institutes of Health (NIH), the world’s largest public funder of biomedical research, examined 149 retracted NIH-funded papers between 1992 and 2012 and reported a direct cost of about $58 million (*4*), but this figure is now outdated. Far more comprehensive accounting is possible today using the Retraction Watch Database (RWD), a curated, regularly updated catalogue of more than 60,000 retraction records across all disciplines (*5*, *6*), and NIH funding metadata, accessible through the agency’s RePORTER system. We integrated the two systems via an automated weekly pipeline and developed a dynamic dashboard that updates continuously as new retractions are cataloged (https://sandovallentisco.shinyapps.io/nih-retractions/). We present here the data for the cost of retracted NIH-funded research as of July 2026 and discuss some implications of these findings.

## Methods

We retrieved the latest version of the Retraction Watch Database (RWD) from the Crossref repository (https://gitlab.com/crossref/retraction-watch-data) and selected entries marked as retractions, excluding expressions of concern or corrections. We also excluded entries classified as ‘Retract and Replace’, and retractions that did not represent a high likelihood of serious error and/or misconduct by the authors (e.g., administrative or publisher errors). For those eligible retracted articles for which RWD identified an institutional affiliation within the U.S., we queried PubMed using the National Center for Biotechnology Information (NCBI) Entrez API to get supplementary funding metadata (grant IDs and number of publications linked to each grant). Finally, to retrieve the funding amount associated with each grant, we used the NIH Research Portfolio Online Reporting Tools Expenditure and Results (RePORTER) tool. Databases from all available fiscal years were downloaded using the ExPORTER tool (see more details about the method in Supplementary Materials). Data and scripts are available at https://github.com/sandovallentisco/nih-retractions-cost.

## Results

### Direct cost to the NIH

As of July 2026, the RWD listed 6,081 retracted articles with at least one U.S. affiliated author. Of these, 1,725 (28.4%) were linked to at least one NIH grant. To calculate the attributed cost per retracted publication, we divided the total grant award by the total number of publications produced under that grant (i.e., attributing a proportional cost to the specific retracted article rather than the entire grant value). The mean attributed cost per retracted NIH-funded article was $155,567 in nominal dollars, $255,087 in 2026 dollars. Summed across all papers, the attributed total direct cost is $268 million in nominal dollars, $440 million in 2026 dollars, about 1% of a single year of NIH budget, but a 7.6-fold increase over the previously published benchmark that covered retractions during 1992-2012 (*4*). The retracted publications were associated with a total of 2,796 distinct NIH grants, i.e., a total of $94.66B in nominal dollars. However, the vast majority of this sum is institutional center and infrastructure funding (e.g., P30 center core grants) rather than money awarded to individual investigators. Considering only investigator-held research, career and fellowship awards (R/K/F-series; see Supplementary materials for the exact activity-code rule) for which the retracted author was the principal investigator, a total amount of $2.63B in nominal dollars or $4.03B in 2026 dollars from 976 grants was associated with retracted papers.

Retracted articles only reflect the work the literature has managed to identify and withdraw, but not the much larger body of flawed or fraudulent work that has yet to be exposed or has been exposed but not yet retracted (*7–9*). Therefore, the $440 million is clearly an underestimate of the wasted resources. The $4.03B can be either an overestimate or an underestimate of the wasted resources. Much of these researchers’ other work may have been without serious flaws; conversely, other work by other researchers who have no retraction yet may have been seriously flawed or even fraudulent.

### Consequences to retracted authors

We tracked the funding trajectories of 1,509 first/last authors for the 3 pre- and post-retraction years. We selected cases in which the first retraction occurred by 2022, so that a full 3-year post-retraction window of funding data was observable for every author in the cohort given the most recent RePORTER fiscal year (2025). Because common author names can match several different NIH investigators, for the 440 authors whose name maps to more than one investigator we kept only the grants of the one whose institution matches their retracted paper (see Supplementary materials for details). We observed some potential decrease in NIH funding for retracted authors. The per-author cohort-wide median annual NIH funding fell from $133,293 in the 3 years pre-retraction to $55,973 in the 3 years post-retraction, a −58.0% change (paired Wilcoxon *V* = 280,530, p = 0.021). This decrease in the median is due to a higher proportion of authors receiving no funding after retraction; the average annual fraction of the cohort holding an active NIH grant fell from 46.7% to 42.3% (McNemar’s test χ^2^(1) = 16.3, p < 0.001). However, the mean remained very similar ($745,851 pre vs $723,275 post; the full per-author funding distribution before and after retraction is shown in the Supplementary Results). The number of unique active grants dropped by 9.9% (paired Wilcoxon *V* = 184,570, p < 0.001). 26.7% of previously funded authors lost all NIH funding in the 3 years after their first retraction.

Changes were more pronounced for authors with more than one retracted paper (n = 332). Their median annual funding fell from $321,648 to $110,200 (−65.7%; paired Wilcoxon *V* = 17,177, p < 0.001), and the mean annual fraction of the cohort holding an active NIH grant fell from 52.5% to 43.5% (McNemar’s test χ^2^ = 12.0, p < 0.001). In contrast to the full cohort, here the mean also decreased substantially ($868,142 pre vs $679,177 post) (See Fig. 1). The number of unique active grants dropped by 19.9% (paired Wilcoxon *V* = 11,520, p < 0.001), and 28.6% of previously funded authors in this subgroup lost all NIH funding in the 3 years after their first retraction.

**Figure 1.**
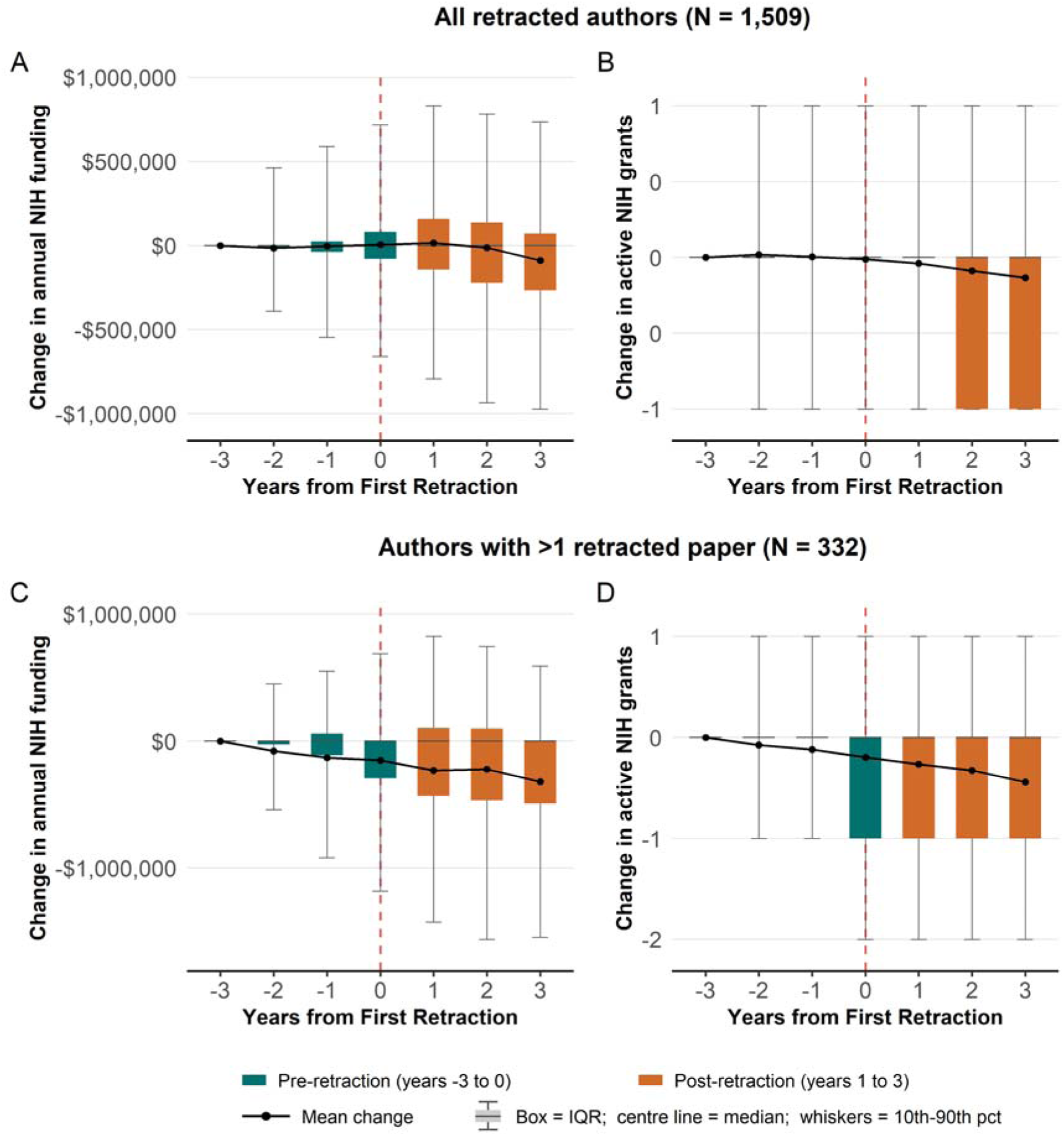
Change in funding for retracted authors compared to year −3. Top row (A, B): the full cohort of 1,509 retracted first/last authors; bottom row (C, D): the 332 authors with more than one retracted paper. (A, C) inflation-adjusted NIH funding; (B, D) count of distinct active NIH grants. Every panel shows the within-author change relative to the baseline year −3 as a boxplot.

### Time to retract NIH-funded articles

Time-to-retraction (see Fig. 2) is markedly longer for NIH-funded papers than for U.S. retractions without NIH funding (6.3 versus 3.13 years on average; Mann-Whitney rank-sum, *W* = 5,388,742, p < 0.001). This gap is largely explained by the different mix of retraction reasons: NIH-funded papers are retracted more often for image-integrity and formal misconduct issues (e.g., data fabrication), which take longer to surface and resolve, whereas non-NIH papers are retracted more often for plagiarism, paper mills, or compromised peer review which may be faster and more definitive to document and retract (see more details in Supplementary Table 2 in Supplementary Materials). The practical implication is that NIH-funded results spend longer in the literature before being retracted, with more time to seed downstream studies, clinical practice, and subsequent funding decisions.

**Figure 2.**
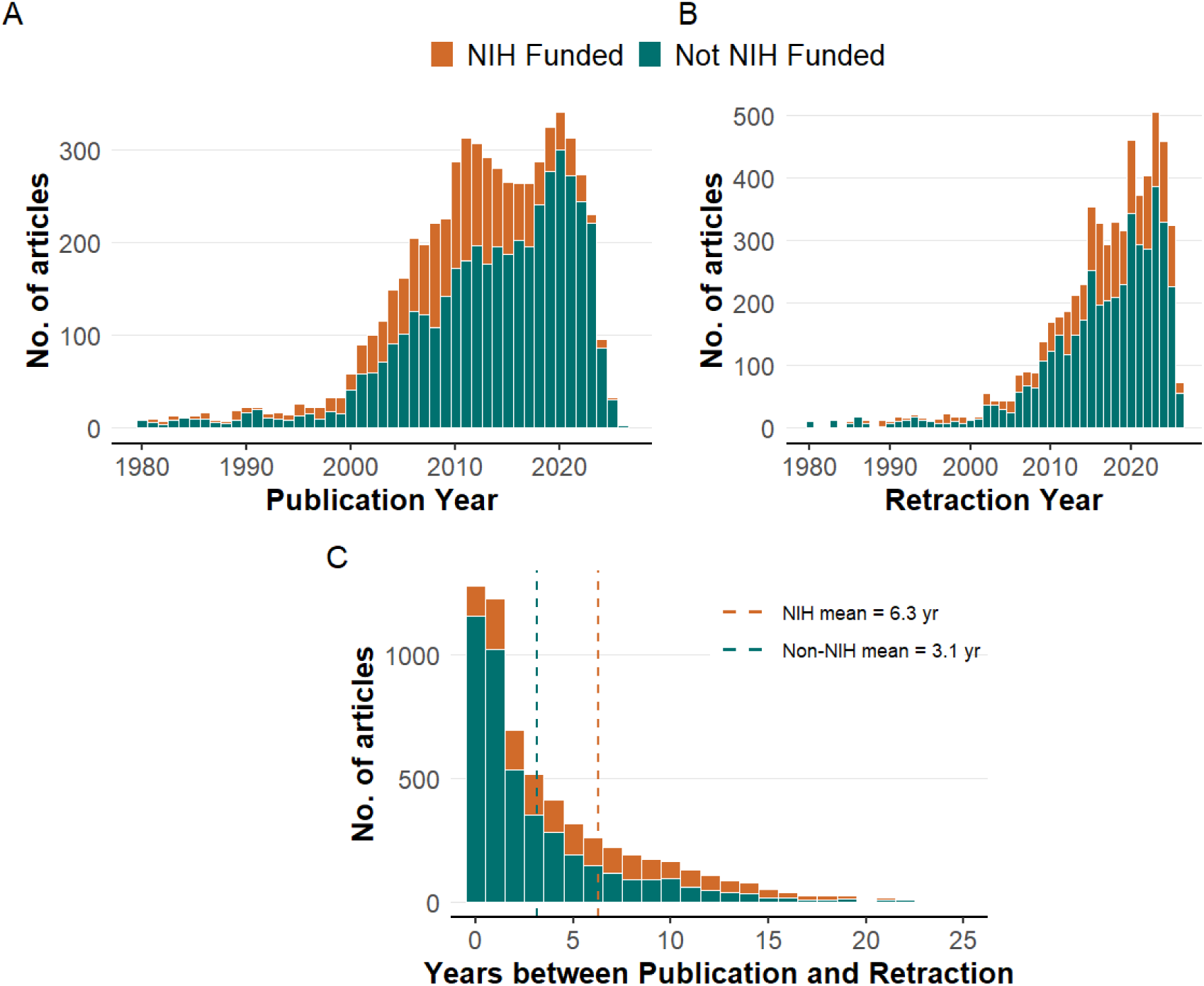
Publication and Retraction Trends. (A) Distribution of publication years. (B) Distribution of retraction years. (C) Lag between publication and retraction; dashed lines mark the mean lag for NIH-funded (orange) and non-NIH (green) papers. Note: For readability, Panels A and B exclude 32 papers published and 12 papers retracted before 1980. Likewise, the X axis of Panel C is capped at 25 years; 15 papers with a lag greater than 25 years are excluded from the plot but included in the means.

## Discussion

According to our estimates, the direct financial costs of NIH-funded retracted only represent a very small portion of the NIH budget. However, far more papers are seriously flawed, misleading, or even fraudulent and never get retracted. Therefore, the wasted resources may be much larger. We also found that investigator-held grants for which the retracted author was the principal investigator were awarded $4.03B in 2026 dollars. Therefore, the footprint of flawed and fraudulent work may relate to a much larger space of research efforts. Furthermore, retraction mechanisms remain slow and inefficient as of today, and they only reflect the visible part of a problem that might be much bigger (*7–9*). Besides, direct cost is also an underestimate of damage since downstream research that built on flawed conclusions or clinical decisions informed by them also entail cost. Evidence synthesis may also be affected (*10*).

Enhancing detection of papers that warrant retraction is not a simple process. Mass-detection tools (image forensics, “tortured-phrase” scanners, statistical anomaly detectors, or emerging AI-content classifiers) may be funded and developed as research infrastructure and applied as pre- and post-publication surveillance. Efficiency and yield of different tools require careful study. Accurate and robust methods to detect fraudulent papers might prevent their publication if applied at the peer review stage or, at least, should enable shorter retraction times, which is especially relevant considering the longer time-to-retraction for NIH-funded studies. Moreover, it should be acknowledged that many retractions reflect honest errors and their authors who detect them and withdraw their flawed work from the published corpus should be congratulated for their diligence and honesty. Creating a culture where honest correction or error is rewarded rather than leading to reputational damage is warranted (*11–13*). Currently, most retractions probably reflect poor research practices and misconduct (*14*), but the balance may change if more honest error retractions are incentivized.

Another important point is that retracted authors experienced a short-term decrease in NIH funding, with the most pronounced declines among authors with more than one retracted article, although a big proportion of previously funded authors still retained at least some funding. One may debate whether and how the NIH might consider enforcing faster and stricter consequences for retracted authors, especially when the reasons for retractions are not due to honest errors, but to serious fraudulent practices such as data fabrication. “Super-retractor” authors with a large number of retractions may account for a sizeable share of total retractions (*15*) and may be a particular group warranting more aggressive measures. Some of these authors may be highly cited and may have a major negative impact in their fields (*16*).

Lastly, considering the fast pace with which the published scientific literature is evolving, static reports quickly become obsolete. The dashboard accompanying this analysis (https://sandovallentisco.shinyapps.io/nih-retractions/) is automatically recomputed weekly from openly available data, so the estimates in this article will continue to refresh after publication. We encourage meta-scientists and policy researchers to also include living, automated analyses of research practices and research outcomes when possible.

## Data Availability

All data and scripts can be found at https://github.com/sandovallentisco/nih-retractions-cost.

## Supplementary materials

Detailed methods and additional analyses can be found at: https://github.com/sandovallentisco/nih-retractions-cost/blob/main/supplementary_materials.pdf

## Disclosures and data availability statement

This study was preregistered on March 23, 2026, on OSF (https://osf.io/guq59/). Deviations from preregistration can be found at https://osf.io/kd8bf/. All data and scripts can be found at https://github.com/sandovallentisco/nih-retractions-cost.

## Supplementary Materials

### Supplementary methods

#### Data extraction and preparation

First, we retrieved the latest version of the Retraction Watch Database (RWD) from the Crossref repository (https://gitlab.com/crossref/retraction-watch-data). We only considered entries marked as retractions and not expressions of concern or corrections. We also excluded entries classified as ‘Retract and Replace’, which indicates that a paper was retracted but subsequently replaced by a corrected version, as this implies the work only contained remediable errors and remains valid in the literature. Additionally, we omitted retractions that did not represent a high likelihood of serious error and/or misconduct by the authors (e.g., administrative or publisher errors), filtering out entries classified as ‘Error by Journal/Publisher’, ‘Duplicate Publication through Error by Journal/Publisher’, or ‘Withdrawn (out of date)’ provided these were not accompanied by additional reasons suggesting author fault.

For those eligible retracted articles for which RWD identified an institutional affiliation within the U.S., we used a custom Python script to automate the retrieval of supplementary funding metadata. This script queried PubMed using the National Center for Biotechnology Information (NCBI) Entrez API to get funding information (grant IDs and and the total number of publications under each grant). We judged the accuracy of PubMed NIH funding information to be good, with very few missing grants (see more details in the next section). In fact, PubMed sometimes captured more grants than were explicitly reported in the article text, which reflects the fact that PubMed also incorporates grant–publication associations from sources beyond the manuscript itself. For example, through the NIH’s My NCBI / My Bibliography system, where investigators link their accounts to eRA Commons and associate their publications with active grants for routine bibliography management and when preparing annual progress reports (RPPRs). Additional grant information might come from publisher deposits, the NIH Manuscript Submission System (NIHMS), and NLM’s own text mining and indexing processes. Together, these mechanisms can capture official funding support even when authors inadvertently omitted the grant numbers from the manuscript’s acknowledgments section. Finally, to retrieve the funding amount associated with a grant, we used the NIH Research Portfolio Online Reporting Tools Expenditure and Results (RePORTER) tool. Databases from all available fiscal years were downloaded using the ExPORTER tool. Grant IDs were searched within these databases, extracting the funding amount associated with each one.

#### Examining the accuracy of PubMed NIH funding information

To assess the accuracy of the retrieved NIH funding data, we qualitatively compared the funding and acknowledgement sections of the first entries (i.e., the most recent entries) from the Retraction Watch Database (RWD) (version retrieved in December 2025) with the information from PubMed. This way, we inspected a total of 123 entries for which there was an associated PubMed ID. Using this sample, we detected that while reported grants from non-NIH funding bodies were frequently missing from the PubMed metadata, NIH grants reported in the articles were correctly included in PubMed in all but two cases (i.e., 2 out of the 45 entries for which some NIH funding was reported). In the first case, all four grants reported in the full text were missing because they were incorrectly disclosed within the Conflict of Interest (COI) section rather than the funding/acknowledgment sections. In the second case, one out of three reported grants was not retrieved, likely missed by the PubMed indexing algorithm due to non-standard reporting formats (it was reported as R 56-139561 instead of R56 HL139561). To mitigate problems arising from misplaced disclosures like in the first case, we also retrieved information about COIs to identify potential grant IDs. In the remaining 78 cases, PubMed did not retrieve any NIH grants because, correctly, no NIH funding had been reported. Nonetheless, because PubMed began indexing COI statements systematically only in 2017 (U.S. National Library of Medicine, 2017), earlier articles may still be subject to missing data.

However, importantly, we also observed in this sample that, in 9 out of the 44 entries for which PubMed identified some NIH grant, the PubMed metadata yielded more NIH grant IDs than those explicitly reported in the article funding/acknowledgement sections. This discrepancy likely happened because PubMed/RePORTER also considers information from authors reported internally, e.g., information submitted to the My NCBI tool to fill in the NIH Research Performance Progress Report (RPPR). Therefore, even if authors failed to disclose funding in the article but submitted this information to other platforms, this information becomes available in PubMed/RePORTER. For this reason, we believe that funding data provided by PubMed is more comprehensive than what is reported in the article. Additionally, we observed minor issues regarding duplicate entries, where the same grant appeared several times with varying formats—e.g., ‘NCI NIH HHS (R01 CA138409)’ and ‘NCI NIH HHS (CA138409)’, or ‘P20RR 15635’ and ‘P20 RR015635’. To prevent double-counting, we implemented an automated cleaning of duplicates based on numeric suffix matching.

After examining this sample, we wanted to determine if PubMed might have poorer retrieval of NIH grants for older cases (although it is also important to note that the number of retractions is much higher in recent publications). Nonetheless, for this reason, but also to further examine the possible issues mentioned above, we randomly selected another 200 entries from the entire sample, ensuring that 100 entries corresponded to articles published before 2005 and the other 100 corresponded to articles published after 2005. After removing the records that lacked an associated PubMed ID, we were finally able to examine 73 and 65 entries from each publication period.

In all cases within this new sample (both before and after 2005), whenever PubMed did not retrieve a grant ID it was because, correctly, no grant ID had been reported in the article either. For the subset of articles published before 2005, PubMed identified at least one NIH grant in 37 entries. Upon inspecting these grants, out of a total of 77 NIH grants IDs that were retrieved by PubMed/74 NIH grants IDs that were explicitly reported in the articles, a total of 6 grants could not be found in RePORTER. Two of these cases not found in RePORTER were published before 1985, which could explain this omission, as RePORTER data only goes back to 1985. In the remaining entries with funding, discrepancies between the number of NIH grant IDs retrieved by PubMed and those reported in the articles were found in only two cases; one because a grant ID was duplicated and in the other cases 2 grant IDs not reported in the text were present in PubMed. After inspecting these cases, we believe it may be a PubMed error since the Fiscal Year was later than the publication of the article. To control these types of potential errors, we excluded grant IDs with fiscal years that post-date the article’s publication.

For the subset of articles published after 2005, PubMed retrieved some NIH funding for 21 entries. For these entries, PubMed retrieved a total of 90 NIH grant IDs, while only 43 were reported in the articles. 24 of these discrepancies were due to duplicate grant IDs, an issue easily resolved by deduplicating as previously mentioned. Most of the remaining 23 discrepancies stem from a case where the article only reports ‘This work was supported by National Institutes of Health grants’ yet PubMed retrieves 14 NIH grant IDs, all of which are linked to one of the authors; we therefore consider this a case where PubMed was more comprehensive than the article itself. Of the remaining 9 discrepant NIH grant IDs, 7 of the grant IDs were also linked to one of the authors, and the other appears to be a PubMed error assigning two placeholder grant IDs (NIA NIH HHS (R99 AG999999); Intramural NIH HHS (Z99 AG999999)), not found in RePORTER. In sum, most discrepancies are due to duplicates that can be resolved or NIH grant IDs that were not reported in the article but are linked to the authors. Spreadsheets with the selected and inspected samples can be found in the ‘Supplementary Materials’ folder of the OSF registration as ‘sample_1.xlsx’ and ‘sample_2.xlsx’ at (https://osf.io/guq59/).

#### Automated “Living” Analysis Pipeline

To ensure the findings remain current (“living analysis”), the entire data retrieval, processing, and deployment pipeline was automated using GitHub Actions. A scheduled workflow is triggered weekly (every Sunday at 06:00 UTC) on a cloud-based environment. The automated pipeline executes the following sequential steps: the workflow initializes a Python 3.11 environment to download the latest raw datasets from the Retraction Watch Database and the NIH ExPORTER. It then executes the core data processing scripts (main.py), interacting with the NCBI Entrez API to fetch updated publication metadata. Once the raw data is cleaned, merged, and analyzed, the workflow automatically commits the newly processed CSV files back to the GitHub repository. This ensures transparent data versioning and maintains a historical record of the dataset over time. In the final stage, the workflow provisions an R 4.3 environment, installs the required dashboard dependencies (e.g., shiny, tidyverse, plotly) directly from the Comprehensive R Archive Network (CRAN), and automatically redeploys the updated Shiny web application to shinyapps.io. The project does not need for manual data updates, guaranteeing that the dashboard consistently reflects the most recent retraction and funding information. The online dashboard is accessible at: https://sandovallentisco.shinyapps.io/nih-retractions/.

#### Estimating the direct cost of NIH-funded retracted articles

We quantify the direct cost in different ways. First, we calculate the *attributed cost* dividing each grant total award by the number of publications produced under that grant and assigns the corresponding share to the retracted paper. This represents the fraction of the grant money that produced a specific retracted paper. We also calculated the money associated with retracted papers directly awarded to researchers in personal grants (i.e., excluding institutional center and infrastructure grants). For this, we first calculate the total award of unique grants (de-duplicating repeated grants to prevent double-counting). Then, to separate grants awarded to an investigator from those awarded to an institution, we use the NIH activity code (e.g., R01, P30, U01). This code is present in the raw ‘Funding_Info’ text (e.g., NCI NIH HHS (R01 CA203737)) in FINAL_Retractions_Costs_and_Pubs.csv. We extract it with a regular expression that captures the three components of a grant number (i.e., activity code, Institute/Center abbreviation, and serial number) allowing the activity code to be either separated from or concatenated with the ID (e.g., U19AI057229, 5P50CA101942). For grants whose code is not recoverable from ‘Funding_Info’, we fall back to the full grant numbers stored in the author–funding file (field ‘Grant_ID’, e.g., K08HL086671, in Author_Funding_Matches.csv). Together these sources assign an activity code to approximately 95% of the unique-grant dollars.

We label a grant as directly awarded to the investigator (“personal”) if its activity code is a research, career-development, or fellowship grant (i.e., codes beginning with R, K or F) or one of the NIH Director’s high-risk personal awards (DP1 Pioneer, DP2 New Innovator, DP5 Early Independence). Within the R-, K- and F-series we exclude mechanisms that are not awarded to fund an individual investigator’s own research, namely institutional career/training mechanisms (K12, KL2), SBIR/STTR awards to companies (R41–R44), and institutional R-series mechanisms; resource grants (R24), education grants (R25), and linked research project awards (RL1). All remaining mechanisms are treated as institutional: center grants (P30, P50, P20, P60, P40, P51, P41), clinical-research centers (M01, UL1), institutional training grants (T-series), shared instrumentation and construction (S10, G-, C06), cooperative agreements (U01, U54, U2C, U19, …; we exclude U01 because, although some fund a single investigator, others are large multi-site consortia that behave more like program or institutional grants), and resource grants from the former National Center for Research Resources (Institute/Center code “RR”). Program-project grants (P01) span several sub-projects and cores across multiple investigators and are treated as institutional here. Grants whose activity code could not be recovered are conservatively not counted as personal.

#### Inflation adjustment

To account for inflation, the attributed cost of each retracted article was adjusted to current-year dollars using the Consumer Price Index (CPI). We matched each article’s publication year to the corresponding historical CPI and calculated an inflation multiplier relative to the most recent available fiscal year. This multiplier was then applied to the raw attributed cost of the publication.

Because a grant award spans several years, there is no single base year to which it can be deflated. For this reason, we assign each grant a representative inflation multiplier equal to the mean of the multipliers of the papers that cite it (each paper’s multiplier is taken from its publication year).

#### Same name correction in the author-consequences analysis

The consequences analysis (Figure 1 of the main text) follows individual authors’ NIH funding across the retraction event. Authors are linked to their NIH grant histories by name, which is reliable for distinctive names but ambiguous for common ones. When a name matches the grant records of several different investigators who happen to share that name, the funding attributed to a retracted author is partly, or entirely, another person’s, which inflates the per-author funding trajectories and can bias the pre/post change.

To address this, we used the NIH ExPORTER project files, in which every project lists the unique person identifiers (PI_IDS) and awardee institution (ORG_NAME) of its principal investigator(s). For each retracted author we collected the distinct PI_IDS that their name maps to across all of their attributed grants. When the name maps to a single PI_ID, every attributed grant belongs to the same person and the link is unambiguous. However, when it maps to more than one PI_ID, the attributed grant history pools two or more different investigators who happen to share that name, a situation we call a “collision”.

To identify whether any of the attributed funding can be tied to the real retracted researcher, we used institutional affiliation. For each attributed grant we compared its awardee institution (ORG_NAME) against the institution listed on the author’s retracted paper. Institutions were matched leniently: each string was upper-cased and reduced to its distinctive tokens (words longer than three characters, excluding generic terms such as “university”, “hospital” or “center”), and a grant was deemed to match the paper when the two shared at least one such token (e.g. the paper affiliation “Dana-Farber Cancer Institute, Harvard Medical School” matches the grant awardee “DANA-FARBER CANCER INSTITUTE”).

We then used this match to clean the attribution at the level of individual grants rather than whole authors. For each collision author we identified the real person’s PI_ID(s) as those holding at least one institution-matching grant, and retained only the grants belonging to that PI_ID, discarding the grants of the same-named others. Because a PI_ID is stable when a researcher changes institution, this keeps the real author’s entire funding trajectory, including grants they later held at other institutions; matching grants by institution alone would instead penalise legitimate institution moves. Authors whose name does not collide keep all of their grants. A collision author with no institution-matching grant has no identifiable real PI_ID, contributes no attributable funding, and therefore leaves the cohort. This grant-level correction is the one used in the main text.

**Supplementary Table 1.** Sensitivity of the author-consequences result to name-collision handling. ‘Change’ is the percentage change in the cohort-wide median per-author annual NIH funding from the three pre-retraction to the three post-retraction years; the Wilcoxon p-value is from the paired signed-rank test; ‘Lost all funding’ is the share of previously funded authors with no NIH funding in the post window. The grant-level PI_ID rule is used in the main text.

| Specification | Authors<br>(N) | Median<br>pre | Median<br>post | Change | Wilcoxon<br>p | Lost all<br>funding |
| --- | --- | --- | --- | --- | --- | --- |
| Full cohort (no correction) | 1,628 | \$145,164 | \$110,200 | -24.1% | 0.219 | 24.7% |
| Grant-level by PI_ID (main analysis) | 1,509 | \$133,293 | \$55,973 | -58.0% | 0.021 | 26.7% |

The two rows contrast the uncorrected cohort with the grant-level PI_ID correction used in the main analysis. Without correction, the attributed funding still pools same-named investigators, whose large, persistent portfolios dilute the decline to about 24%. The grant-level PI_ID rule isolates each real author’s own grants—for an author whose name matches several investigators, keeping only the grants of the one whose awardee institution matches their retracted paper—after which the cohort-wide median annual funding falls by about 58% (paired Wilcoxon p = 0.021), a decline that, unlike the uncorrected cohort, reaches statistical significance. The magnitude of the decline is thus sensitive to how name collisions are handled, and the headline figure should be read with that caveat; the direction, i.e., a substantial post-retraction decline in the funding genuinely attributable to retracted authors, holds once the attribution is cleaned.

### Supplementary Results

#### Distribution of per-author funding

The corrected cohort-wide median falls by about 58%, but this reflects mainly an increase in the share of authors with no NIH funding rather than a uniform reduction across the cohort. The figure below shows the per-author annual funding before and after retraction. For the full cohort the two distributions, and their means, are very similar; the change is concentrated in the median. For authors with more than one retracted paper, the whole distribution, including the mean, shifts downward.

**Supplementary Figure 1.**
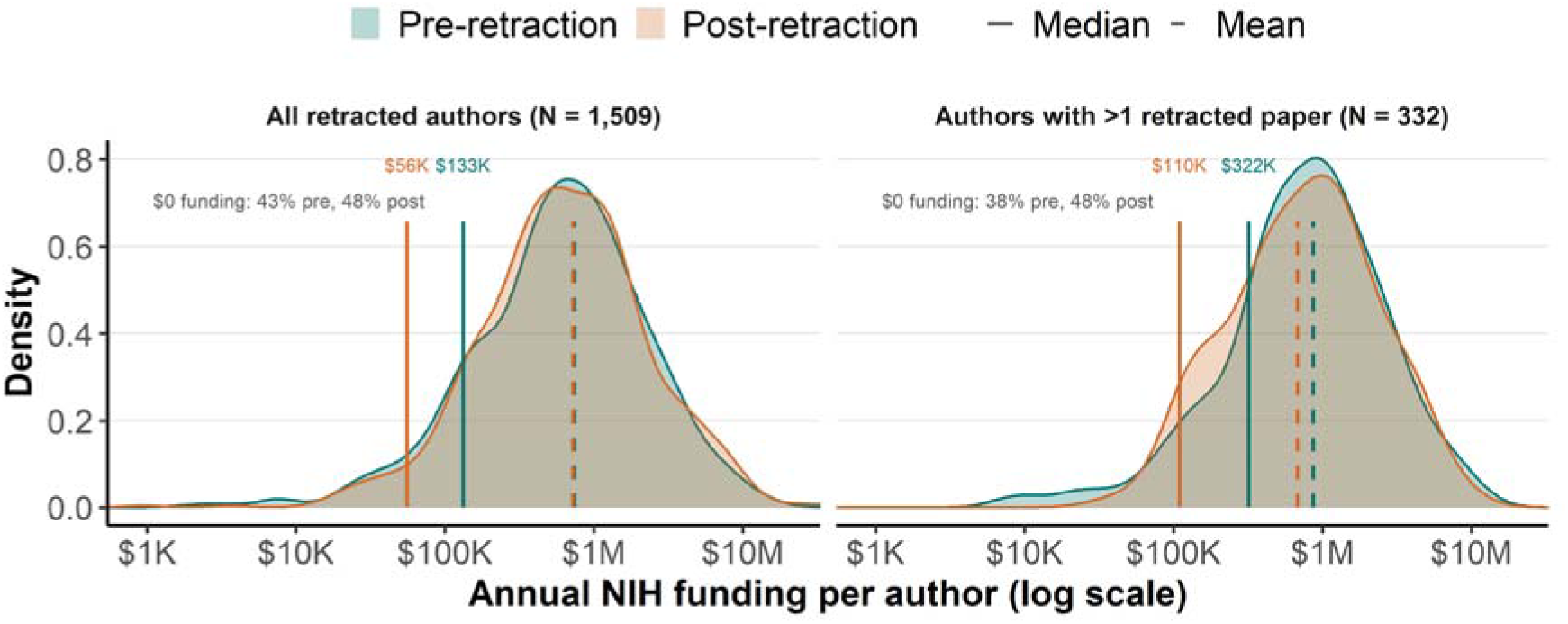
Distribution of per-author annual NIH funding in the three years before (green) and after (orange) retraction, after the grant-level collision correction, for authors with non-zero funding (kernel density on a log scale; zero-funding authors are omitted from the curve but their share is annotated in each panel).

#### Top retraction reasons by funding source and time-to-retraction

NIH-funded and non-NIH-funded retracted papers differ systematically in why they are retracted, and these reasons differ in how quickly they are acted on, which accounts for much of the longer time-to-retraction of NIH-funded papers reported in the main analysis. NIH-funded papers are concentrated in data- and image-fabrication/manipulation and in formal misconduct investigations, which surface late (through image forensics and institutional or ORI investigations) and take years to resolve; non-NIH papers are more often retracted for plagiarism and paper mills, which are acted on much faster. The table below shows, for each broad reason category, the share of papers in each funding group whose retraction notice lists such a reason, together with the median time-to-retraction for papers with that reason.

**Supplementary Figure 2.**
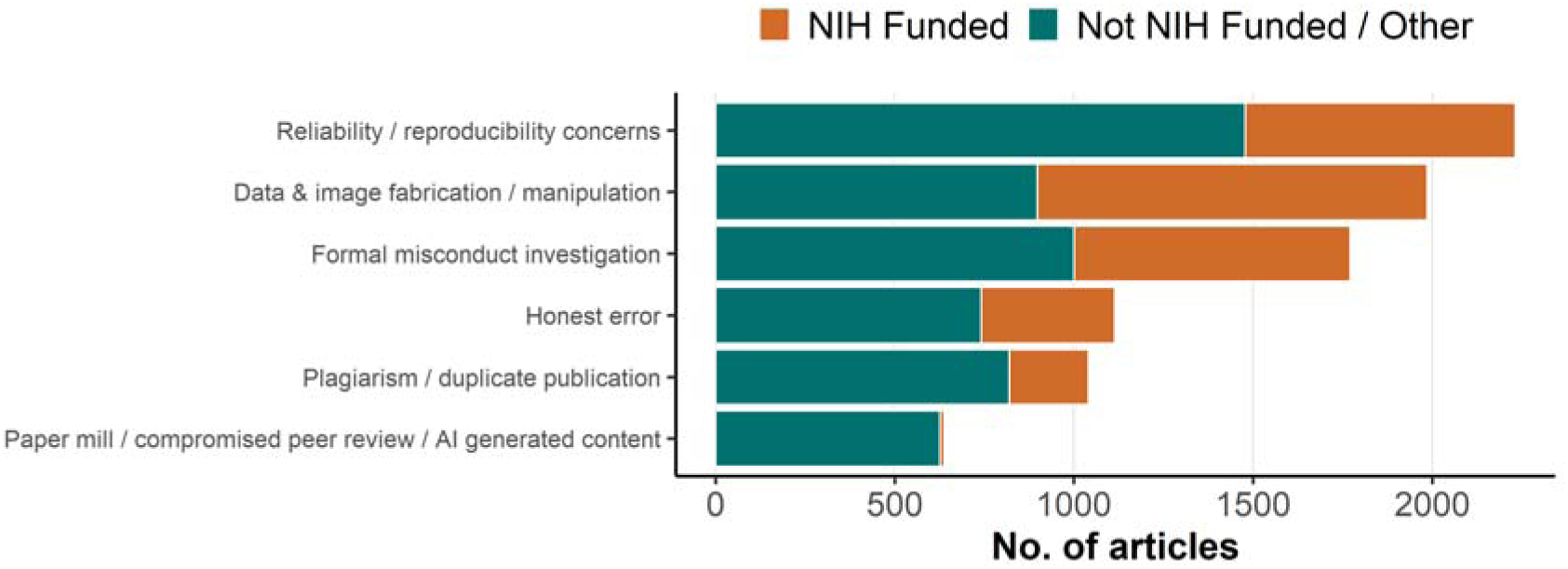
Retraction reasons by funding source, grouped into broad categories.

**Supplementary Table 2.** Retraction reasons by funding source and their median time-to-retraction.

| Retraction reason | % of NIH-funded | % of non-NIH-funded | Median lag (years) |
| --- | --- | --- | --- |
| Data & image fabrication / manipulation | 63.1% | 20.6% | 6 |
| Formal misconduct investigation | 44.7% | 23.0% | 5 |
| Reliability / reproducibility concerns | 43.8% | 33.9% | 3 |
| Plagiarism / duplicate publication | 12.8% | 18.8% | 2 |
| Honest error | 21.6% | 17.0% | 2 |
| Paper mill / compromised peer review / AI generated content | 0.7% | 14.4% | 1 |

The categories are our own aggregations of the raw Retraction Watch Database reason labels, grouping the most frequent substantive reasons into six broad mechanisms. Because a single retraction notice can list several labels, the categories are not mutually exclusive; and because the database uses many fine-grained labels, the categories do not capture every label; some substantive labels (e.g., research-ethics and consent, authorship, or referencing/attribution issues) fall outside the six categories. Procedural and administrative reasons (e.g., Investigation by Journal/Publisher) and notice-status labels (Notice – Limited or No Information; Date of Article and/or Notice Unknown; Upgrade/Update of Prior Notice(s); Removed; Notice – Unable to Access via current resources) are not included, as they describe the retraction process or notice status rather than a substantive cause.

Each category groups the following raw labels — *Data & image fabrication / manipulation*: Duplication of/in Image; Manipulation of Images; Falsification/Fabrication of Image; Concerns/Issues about Image; Plagiarism of Image; Unreliable Image; Falsification/Fabrication of Data; Falsification/Fabrication of Results. *Formal misconduct investigation*: Investigation by Company/Institution; Investigation by ORI; Investigation by Third Party; Misconduct - Official Investigation(s) and/or Finding(s); Misconduct by Author. *Reliability / reproducibility concerns*: Unreliable Results and/or Conclusions; Unreliable Data; Concerns/Issues about Data; Concerns/Issues about Results and/or Conclusions; Results Not Reproducible; Original Data and/or Images not Provided and/or not Available. *Plagiarism / duplicate publication*: Plagiarism of/in Article; Plagiarism of Text; Plagiarism of Data; Duplication of/in Article; Duplication of Text; Euphemisms for Plagiarism; Euphemisms for Duplication; Duplication of Data. *Honest error*: Error in Data; Error in Analyses; Error in Results and/or Conclusions; Error in Methods; Error in Image. *Paper mill / compromised peer review / AI generated content*: Compromised Peer Review; Concerns/Issues about Peer Review; Paper Mill; False/Forged Authorship; False/Forged Affiliation; Computer-Aided Content or Computer-Generated Content.

Unlike the other categories, “Formal misconduct investigation” does not describe a specific defect of the paper but rather the procedural pathway through which the retraction was reached (i.e., a formal investigation by an institution, the ORI, or a third party). Accordingly, 95% of papers in this category also carry a label from at least one other category (most often data/image fabrication or manipulation, or reliability/reproducibility concerns). Because NIH-funded papers are routed through these formal investigations far more often than non-NIH papers, this slow institutional/ORI pathway accounts for a substantial part of their longer time-to-retraction.

#### Scientific domains and publishers of U.S. retracted articles

**Supplementary Figure 3.**
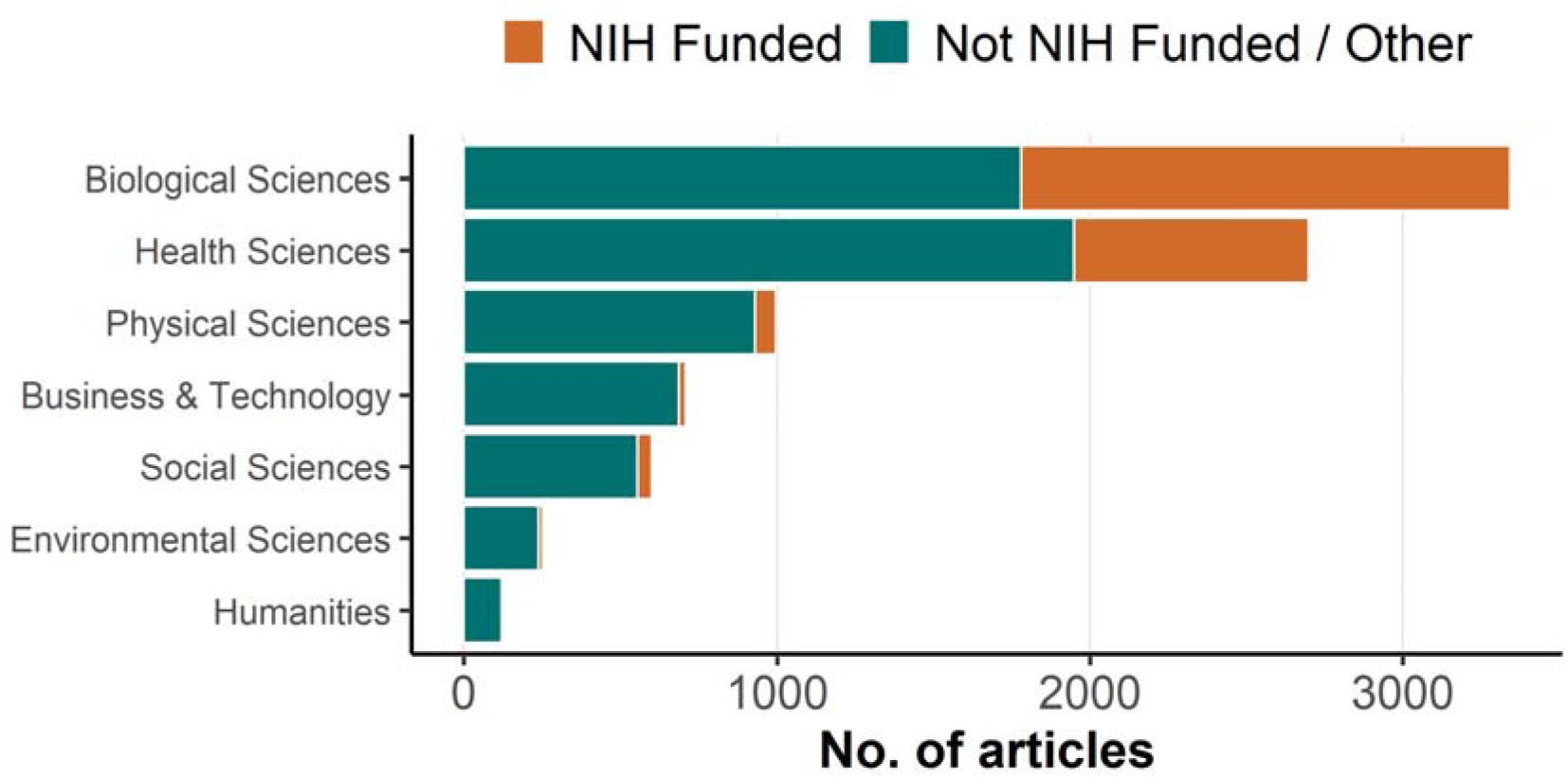
Scientific domains by funding. Note. A single paper can have multiple subjects, but each broad category is counted at most once per paper.

## References

1. I. Oransky, A. Marcus, Why write a blog about retractions? (2010). https://retractionwatch.com/2010/08/03/why-write-a-blog-about-retractions/.

2. P. Sebo, M. Sebo, Geographical Disparities in Research Misconduct: Analyzing Retraction Patterns by Country. Journal of Medical Internet Research 27, e65775 (2025).

3. R. G. Steen, A. Casadevall, F. C. Fang, Why Has the Number of Scientific Retractions Increased? (American Psychological Association, Washington, DC, US, 2016) *Methodological issues and strategies in clinical research, 4th ed*.

4. A. M. Stern, A. Casadevall, R. G. Steen, F. C. Fang, Financial costs and personal consequences of research misconduct resulting in retracted publications. eLife 3, e02956 (2014).

5. I. Oransky, The retraction watch database becomes completely open and RW becomes far more sustainable (2023). https://retractionwatch.com/2023/09/12/the-retraction-watch-database-becomes-completely-open-and-rw-becomes-far-more-sustainable/.

6. P. Sebo, M. Sebo, Comparing the performance of retraction watch database, PubMed, and web of science in identifying retracted publications in medicine. Accountability in Research 0, 1–25 (2025).

7. M. Naddaf, E. Quill, Hallucinated citations are polluting the scientific literature. What can be done? Nature 652, 26–29 (2026).

8. B. Scancar, J. A. Byrne, D. Causeur, A. G. Barnett, Machine learning based screening of potential paper mill publications in cancer research: methodological and cross sectional study. BMJ 392, e087581 (2026).

9. M. Topaz, N. Roguin, P. Gupta, Z. Zhang, L.-M. Peltonen, Fabricated citations: An audit across 2·5 million biomedical papers. The Lancet 407, 1779–1781 (2026).

10. C. Bakker, S. Boughton, C. M. Faggion, D. Fanelli, K. Kaiser, J. Schneider, Reducing the residue of retractions in evidence synthesis: Ways to minimise inappropriate citation and use of retracted data. BMJ Evidence-Based Medicine 29, 121–126 (2024).

11. D. Fanelli, J. P. A. Ioannidis, S. Goodman, Improving the integrity of published science: An expanded taxonomy of retractions and corrections. European Journal of Clinical Investigation 48, e12898 (2018).

12. M. Hosseini, M. Hilhorst, I. de Beaufort, D. Fanelli, Doing the right thing: A qualitative investigation of retractions due to unintentional error. Science and Engineering Ethics 24, 189–206 (2018).

13. D. Fanelli, Set up a ‘self-retraction’ system for honest errors. Nature 531, 415 (2016).

14. S. Y. Hwang, D. K. Yon, S. W. Lee, M. S. Kim, J. Y. Kim, L. Smith, A. Koyanagi, M. Solmi, A. F. Carvalho, E. Kim, J. I. Shin, J. P. A. Ioannidis, Causes for retraction in the biomedical literature: A systematic review of studies of retraction notices. Journal of Korean Medical Science 38, e333 (2023).

15. C. Lyu, M. Matbouriahi, F. Naudet, J. P. A. Ioannidis, I. A. Cristea, Retracted randomized clinical trials from superretractors and top-cited scientists with multiple retractions. JAMA Network Open 9, e267424 (2026).

16. J. P. A. Ioannidis, A. M. Pezzullo, A. Cristiano, S. Boccia, J. Baas, Linking citation and retraction data reveals the demographics of scientific retractions among highly cited authors. PLOS Biology 23, e3002999 (2025).

